# Role of Flipped Classroom Method in Short and Long Term Retention in Anatomy

**DOI:** 10.1101/2020.08.01.20166280

**Authors:** Payal Arvind Kasat, Vishwajit Deshmukh, Gayatri Muthiyan, T.S. Gugapriya, Aaditya Tarnekar, Bharat Sontakke, Smita Sorte

## Abstract

**Introduction:** Medical education is changing towards more flexible, effective, active and student-centered teaching strategies that reduces the limitations of traditional methods of education. Recently, the flipped classroom method has been suggested to support this transition. However, research on the use of flipped classroom method in medical education is at its early stage and little is known about its effect on students learning in relation to short and long-term retention of the topics.

**Aims:** The present paper aims to study the comparative effect of traditional and flipped classroom method on short-term and long-term memories of first MBBS students in Anatomy with the aid of technology to promote learning.

**Materials and methods:** 50 first year MBBS students were subjected to traditional and flipped classroom module separately. Immediate assessments were done at the completion of the module. Followed by a gap of 2 months, the students were again assessed on the content taught in the module as a part of formative assessment. The data so obtained was compared and analyzed statistically.

**Result:** The assessment scores showed differences between the two methods of teaching in short as well as long term. The flipped classroom method was observed to have significant long-term retention which was evident by assessment scores.

**Conclusion:** The study concludes that flipped classroom method serves as an advantageous tool and motivating factor for effective learning, understanding and retention of conceptual and factual anatomical content.

## Introduction

From historical times, teaching was in the form of “The sage on the stage”. Traditional classroom portrays teachers as content providers and students as receivers. Thus, in traditional classroom setting, students have been made passive in the process of knowledge acquisition [1]. In contrast to traditional methods of teaching anatomy, flipped classroom teaching (FCT) is a pedagogical intervention which is beneficial for students. It is defined as a type of blended teaching-learning method in which students are introduced to content of subject at home in the form of presentation, pictures, videos, notes etc. and students will practice concepts and applied aspects during the flipped sessions [2].

Thus, the didactic lectures, which are usually taking place during the face-to-face time, are prerecorded and made available for students before the flipped classroom. During flipped class they perform learning activities such as exercises, projects, or discussions. So, they can gain knowledge and conceptualize them to attain higher levels of Bloom’s taxonomy. A flipped classroom inverts the typical cycle of content acquisition and application. The class time and conventional homework time are exchanged. Homework is done first and class time is effectively used for personalized learning which serves as a new paradigm in medical education. [3,4]. (Figure 1).

**Fig. 1.**
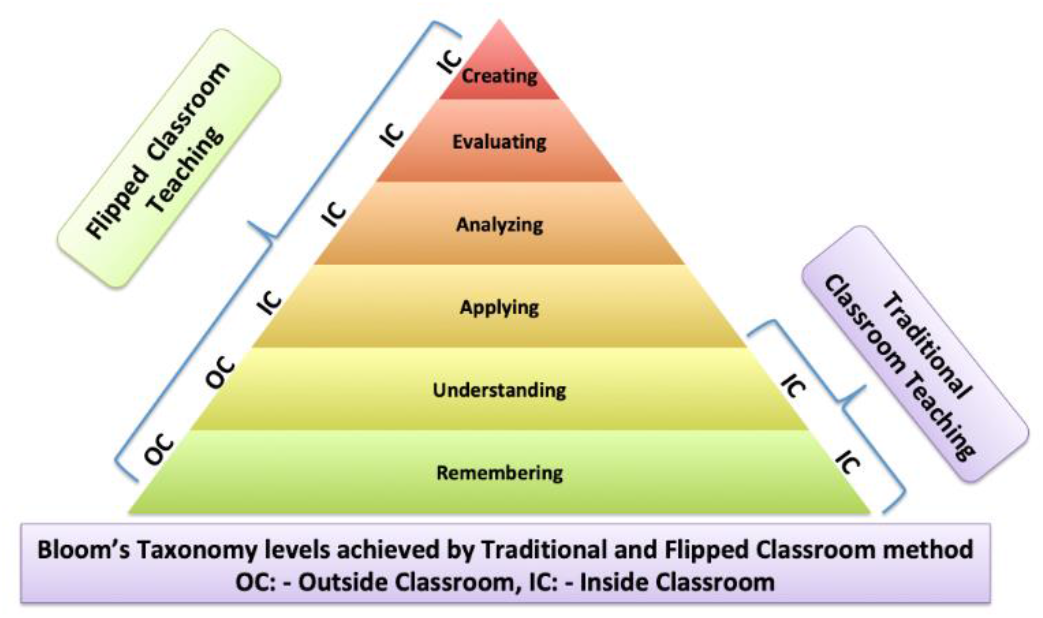
Illustration showing Bloom’s taxonomy levels achieved by Traditional and Flipped classroom method

Recent studies of learner’s perception of the flipped classroom in health professionals found an overwhelmingly positive response from students after attending the flipped classroom. Students also highly appreciated the regular formation of small groups for discussion-based activities, which are face to face that help to change their attitude towards learning, furnish motivation to learn, increase their level of engagement and most important increases interest of the subject per se. Students expressed high levels of satisfaction with pre-class videos and presentation because they have unrestricted access to pre-recorded video lectures at any time and place, according to the student’s convenience and at their own pace [5, 6]. (Figure 2)

**Fig. 2.**
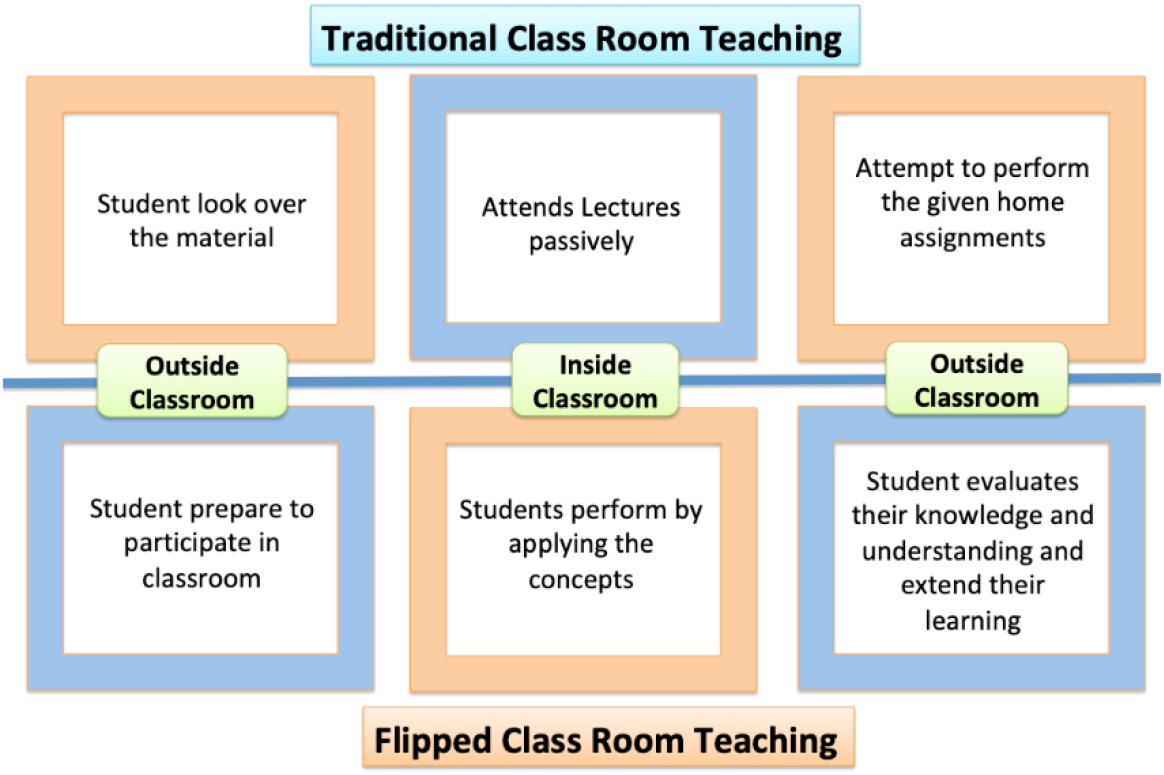
Illustration showing differences in Traditional and Flipped Classroom teaching

## 2. Aims

The purpose of this research was to study the comparative effect of traditional and flipped classroom method on short-term and long-term memories of first MBBS students in Anatomy with the aid of technology to promote learning.

## 3. Objectives

The objectives of the study were:

1. To obtain assessment scores immediately after completion of traditional teaching module.
2. To obtain assessment scores immediately after completion of flipped teaching module.
3. To obtain assessment scores 2 months after completion of traditional teaching module.
4. To obtain assessment scores 2 months after completion of flipped teaching module.
5. To compare the effect of these modules on the memories of student.
6. To obtain students feedback regarding flipped classroom teaching.

## 4. Materials and Methods

In the present study, fifty first year undergraduates were included. The study was conducted in the Department of Anatomy of a teaching institute. They were subjected to traditional and flipped modules separately during Anatomy teaching hours. During the traditional module, routine large group didactic sessions were conducted.

In flipped module, students were provided with pre-recorded self-made videos, power point presentations and pdf documents from reference textbooks etc. one week prior to the scheduled sessions on WhatsApp group. They were instructed to:

a. Hear the prerecorded videos prepared by faculty / facilitator.
b. Read the study material (textbook or notes or ppts).
c. Make mind-maps or notes for the topic.
d. Practice questions given for the topic.
e. Jot down queries that arise during pre-flipped session.

Following which during the two hours flipped classroom sessions, the students were divided into small groups of five. Each group had a facilitator assigned. At the beginning the facilitator would provide case-based scenarios for engaging students in higher orders of critical thinking and problem solving. The facilitator would be observing the discussion passively and will intervene only at the end for giving feedback, clarifying misconceptions, help understand challenging concepts, resolving doubts and concluding the session. At the end the facilitator would give personalized assignments to the students.

During these discussions, the students were expected to:

a. Systematically present the topic (peer-teaching).
b. Answer the queries of colleagues based on their understanding (cooperative learning).
c. Seek explanation from the facilitator for any queries left unsolved (customized learning).

Immediate assessments were done at the end of each module. After a gap of 2 months, the students were assessed on the content taught in the module as a part of formative assessment. The data so obtained was compared and analyzed statistically. Feedback from students about teaching methods was also taken. The study was approved by the Institutional Ethics Committee (IEC). The results obtained were recorded and tabulated. The different parameters recorded were:

1. Assessment scores immediately after completion of traditional teaching module.
2. Assessment scores immediately after completion of flipped teaching module.
3. Assessment scores 2 months after completion of traditional teaching module.
4. Assessment scores 2 months after completion of flipped teaching module.

The data was statistically analyzed for the purpose of comparison and correlation by calculating the mean, range and standard deviation. t-test with two-tailed distribution was applied for comparison of values of different parameters. p-value< 0.05 was considered as statistically significant. Spreadsheets were used to tabulate values.

## 5. Results

1. The difference between the assessment scores for short term between flipped classroom module versus traditional classroom module was statistically significant with p-value <0.00001. (Table 1, Figure 5)
2. The assessment scores for long-term had increased for both the teaching modules when compared with respective assessment scores for short term. (Table1, Figure 3, Figure 4)
3. The assessment scores of flipped classroom module are more compared to the assessment scores of traditional classroom module in short as well as long term. (Table 1, Figure 5, Figure 6)
4. There is difference between the long-term assessment scores for flipped classroom module versus traditional classroom, however it is not statistically significant with p-value <0.095.
5. The assessment scores for long term are almost uniformly elevated for flipped classroom module. (Figure 4)
6. The difference between short- and long-term assessment scores for traditional module is more than the same for the flipped module. (Table 1)
7. The number of students who gained more than 50% scores in short term assessment for flipped module is significantly more when compared with the similar group for traditional module. (Table 2)
8. The number of students who gained less than 35% scores in short term assessment for flipped module is significantly less when compared with the similar group for traditional module. (Table 2)

**Fig. 3.**
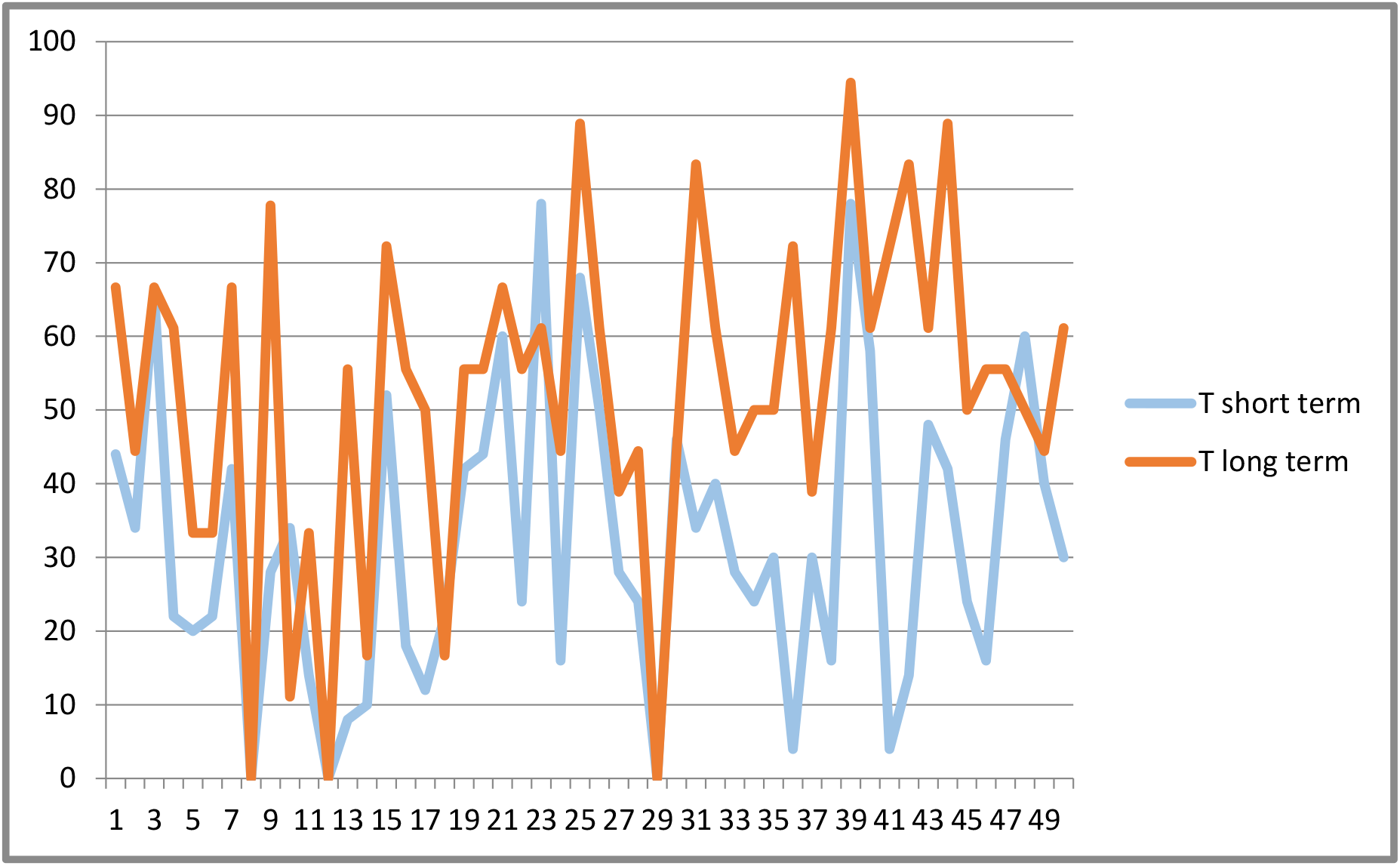
Illustration showing graph of short and long-term assessment scores of students following traditional module

**Fig. 4.**
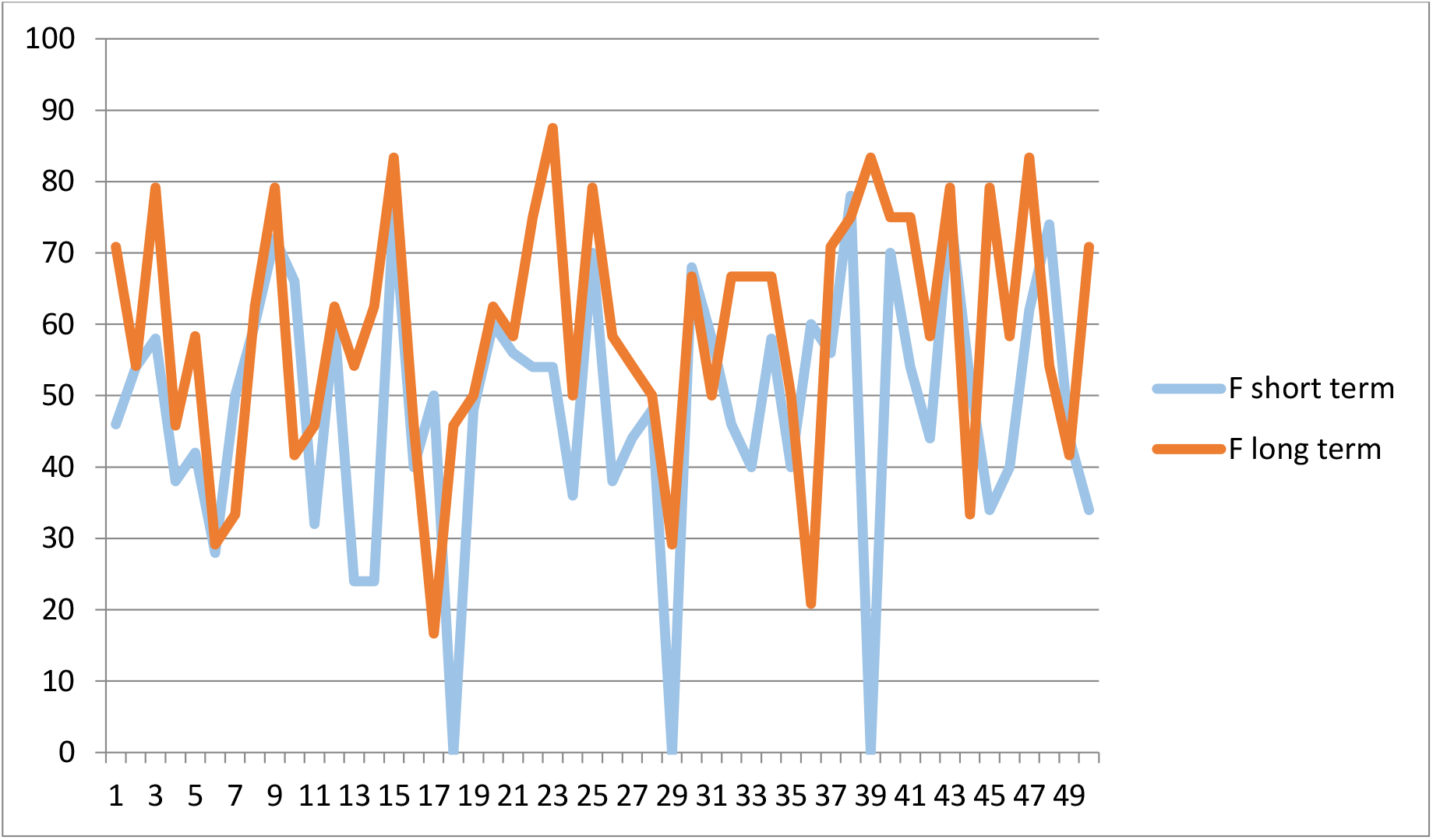
Illustration showing graph of short and long-term assessment scores of students following flipped module

**Fig. 5.**
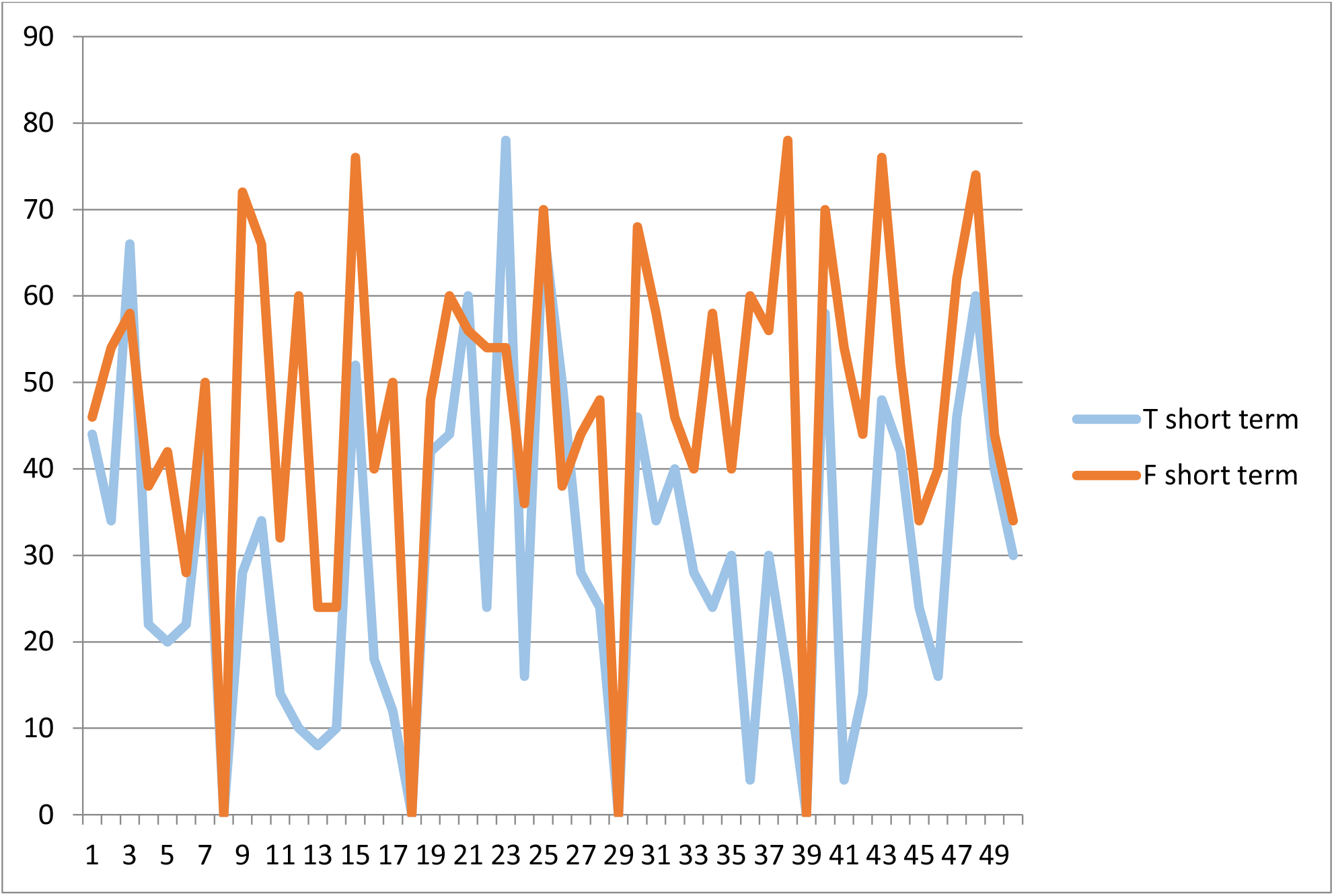
Illustration showing a comparative graph of short-term assessment scores of students following traditional and flipped module

**Fig. 6.**
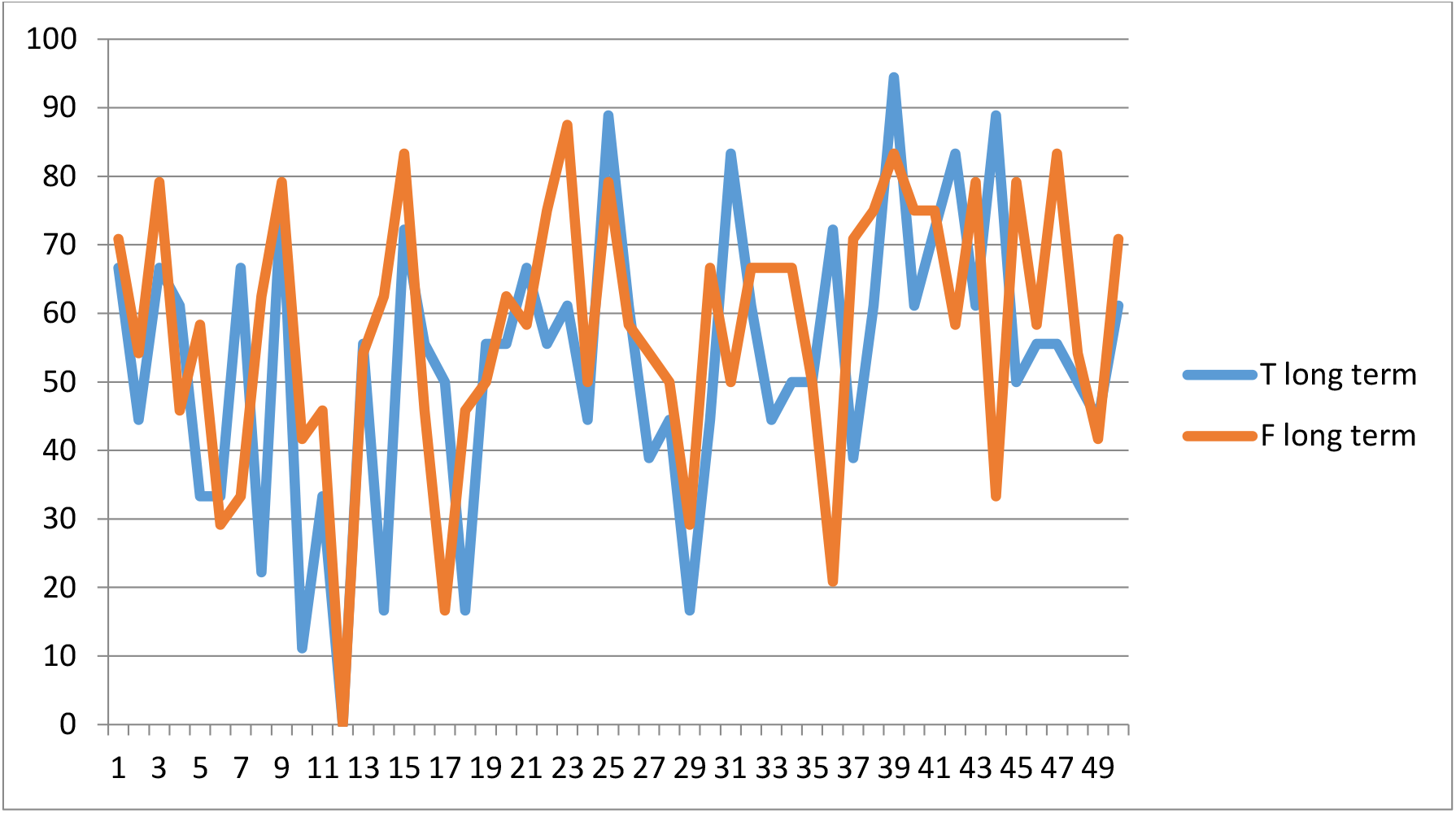
Illustration showing a comparative graph of long-term assessment scores of students following traditional and flipped module

**Table 1:**
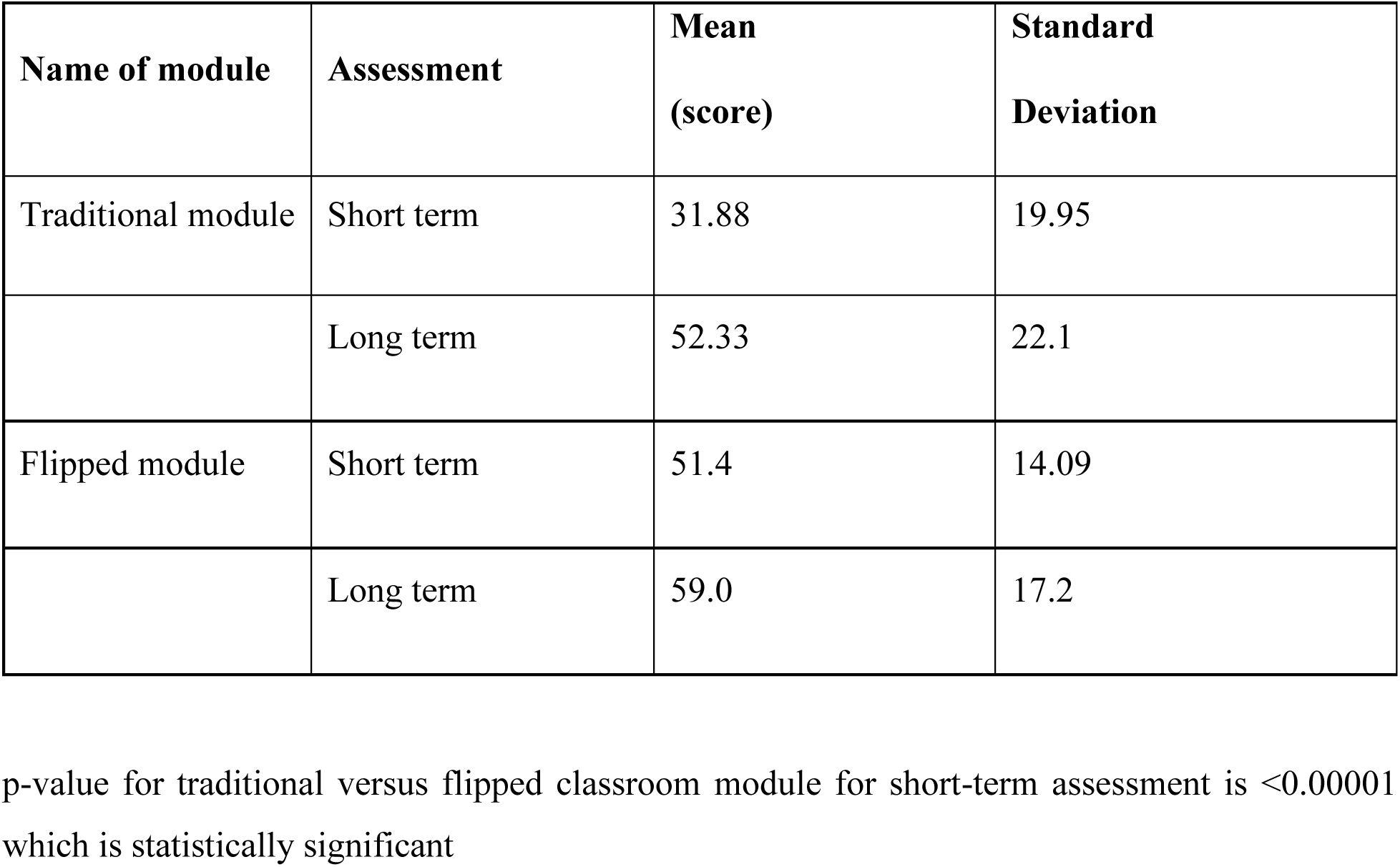
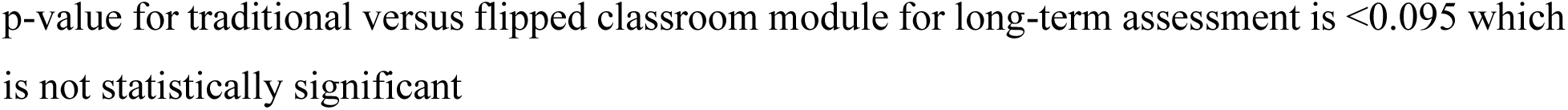
Short-term and long-term scores for traditional and flipped module.

| Name of module | Assessment | Mean<br>(score) | Standard<br>Deviation |
| --- | --- | --- | --- |
| Traditional module | Short term | 31.88 | 19.95 |
|  | Long term | 52.33 | 22.1 |
| Flipped module | Short term | 51.4 | 14.09 |
|  | Long term | 59.0 | 17.2 |

p-value for traditional versus flipped classroom module for short-term assessment is $<0.00001$ which is statistically significant
p-value for traditional versus flipped classroom module for long-term assessment is <0.095 which is not statistically significant

**Table 2:**
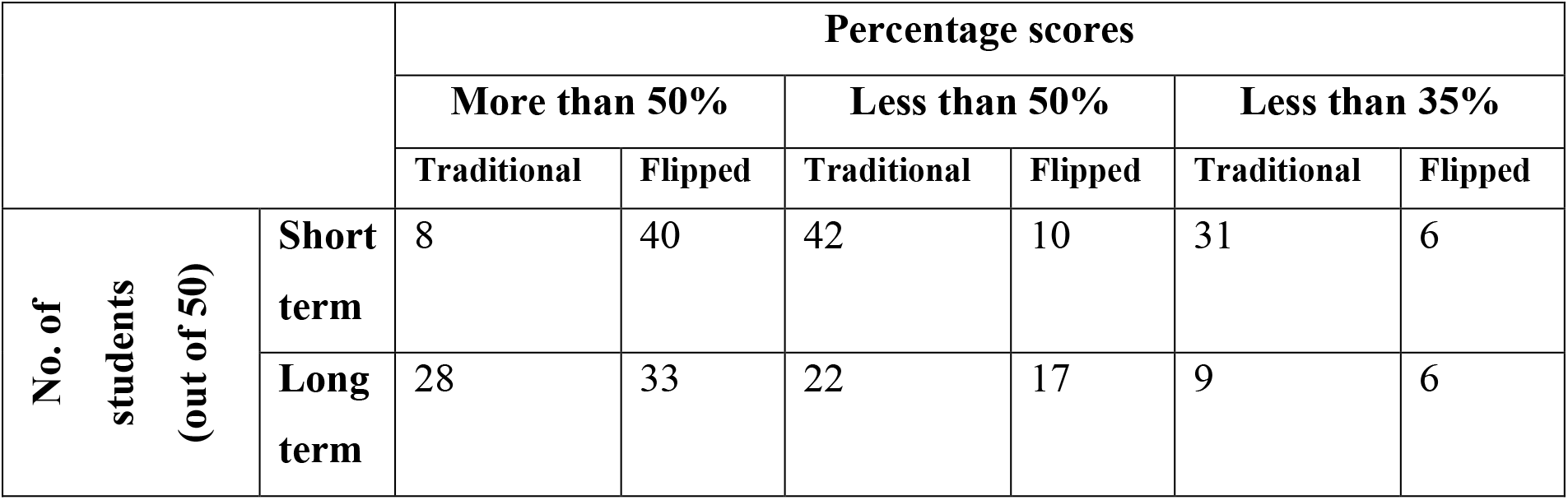
Count of Students based on the percentage scores for flipped and traditional modules.

|  |  | Percentage scores |  |  |  |  |  |
| --- | --- | --- | --- | --- | --- | --- | --- |
|  |  | More than 50% |  | Less than 50% |  | Less than 35% |  |
|  |  | Traditional | Flipped | Traditional | Flipped | Traditional | Flipped |
| **No. of students (out of 50)** | **Short term** | 8 | 40 | 42 | 10 | 31 | 6 |
|  | **Long term** | 28 | 33 | 22 | 17 | 9 | 6 |

Thematic analysis of the students’ feedback

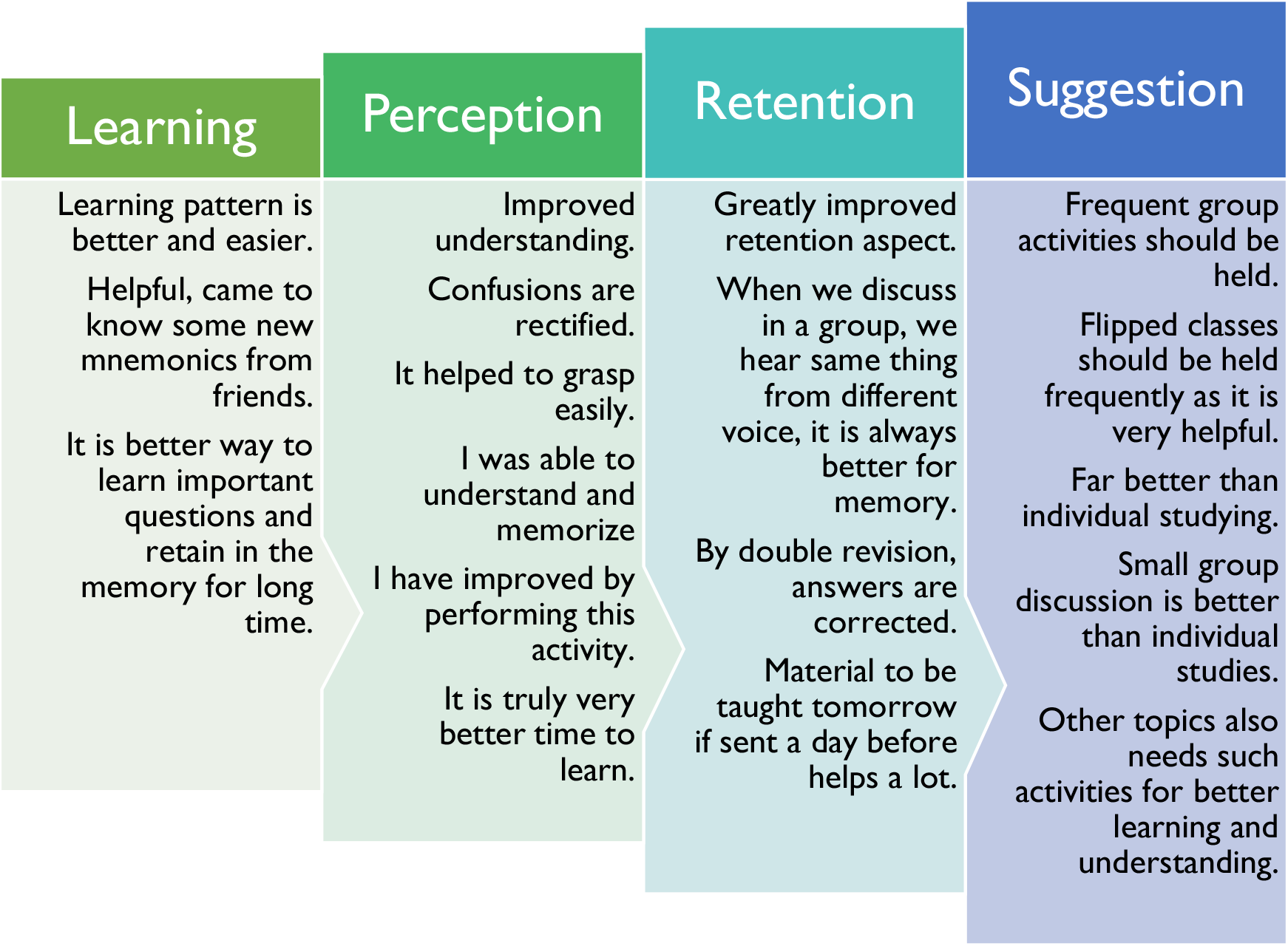

## 5. Discussion

In this study, the effect of flipped classroom technique on the memory retention of the undergraduates was evaluated. In terms of assessment of learning outcomes, researchers have demonstrated higher exam grades for students using a flipped classroom approach as compared to students learning through traditional methods [7]. The improvement in the assessment scores explains success of flipped classroom teaching techniques. It is obvious by the impact on the generation effect and the testing effect pertaining to this study. The generation effect is defined as the ability to regenerate the knowledge at a higher rate than the knowledge given to them during the routine reading. This is more in flipped than traditional method as is evident from the statistically significant difference in the short-term assessment scores (Figure 5, table 1) [8].

The testing effect studies the impact on the students’ long-term memory. This has improved in both the methods. The long-term assessment scores for flipped module are relatively more than for traditional module. However, the difference between long term assessment scores and the short-term assessment scores for traditional module is much more than that for the flipped module. This is because the students had gained very low short-term assessment scores for the traditional module. This greater difference can possibly be explained by the fact that students tend to read the confined syllabus just before the exam. So, their scores for formative assessment examinations are better. However, if the students fail to revise the portion covered in flipped/ traditional class then it is likely that the score may decrease unless the student has good retention capacity. The ability of each individual students to go in depth of the topic will also determine their assessment scores. Hence, we suggest surprise assessment for these topics covered in flipped classroom. This will also give a fair idea about the actual impact of these sessions on long term retention [8].

Also, the difficulty to write theoretical answers based on the flipped classroom module could be another confounding factor. The flipped classroom module emphasizes more on understanding the concepts than on writing theoretical answers, which are necessarily present in the design of the formative examinations. When the assignments for the flipped module are designed to engage the students in answering theoretical questions, they will outperform in final test as compared to those who study only for the final test [8].

It is substantiated by students that after having gone through the study material provided prior, when they discuss in a group and hear the same from fellow colleague, then it helps them to remember and retain for long time. This finding is in accordance to the Ebbinghan’s Forgetting curve, which states that there is direct correlation between memory and time. Generally, forgetting is a rapid process. But to remember the things, repeated practice is required. Experiments have shown that if one memorizes repeatedly within an hour, one will remember the things for a day. If one memorizes one day later, one will remember for one week [9].

Students like the flipped classroom teaching and prefer it over learning alone individually. Students take responsibility and control of own learning in terms of pace of study, mastery of content and for coming to class prepared. Each student has his/her own method of learning and during flipped sessions they interact and become aware of new techniques of learning for the same topic like mnemonics, sharing of personal experience etc. This is beneficial in building concepts and in development of critical thinking. Also, they are more receptive and do not hesitate to clear their doubts during such sessions. While actively participating in the discussions during the flipped classroom the students can reach higher levels in Bloom’s taxonomy, such as application, analysis and synthesis of knowledge and attain leadership quality and ability to work in team. This is in conformity with the previous reports which compares three different instructional strategies in an information systems spreadsheet course. The study showed that students attending the flipped classroom course were more satisfied with the learning environment compared to the other treatment groups [4]. Students expressed that flipped classroom is very useful and that it should be arranged more often. This is in line with the previous study where analysis of pharmacy students’ experiences of flipped classroom courses revealed that students who learned through a flipped classroom approach considered themselves more engaged than students attending traditional courses [10].

According to one of the studies performed on the graduates in the internal medicine department, residents interview expressed significant concerns about the time and motivation to watch the required videos outside of scheduled teaching sessions [11]. When the students find it difficult to put in required preparatory time and effort for viewing the precourse material, in that case the students will join the flipped classroom passively [12]. Thus, the limitation for the study was ensuring whether students coming for the flipped classroom sessions were going through the material beforehand or they joined the discussion passively. We suggest that along with the study material we also give a list of questions pertaining to the topic which students can prepare. At the start of the flipped classroom, a quiz in the form of MCQ test, pictorial questions, word search, cross word puzzle etc. can be held which will act as a major factor to increase the student’s interest in grasping the content. The quiz questions will be prepared by the facilitator beforehand and will be used to assess the students concerning the material given for the flipped classroom. Students are likely to be able to solve many of the questions. However, in case they are unable to do so, still they are determined to get answers to those questions during the flipped classroom sessions. They will be very much alert and actively involved. Also, this will improve the assessment scores post-flipped classroom sessions as students have put in their efforts for learning and understanding the topic under discussion.

Quiz at the start of the flipped classroom is beneficial for students to recall the knowledge gained by the material provided prior hand, as prior hand knowledge has been considered as an important factor influencing learning. If students can retrieve the knowledge gained from the preclass material, it makes the information more rigid in the form of memory, which can be applied in future occasions. Also, the instructor will identify any sort of misconceptions related to the topics from the preclass material. Based on the concepts gained, the instructor can provide remedial actions if required in the form of reviewing the material again. Lastly, it may act as a strong motivator for the students to watch and learn the preclass material before joining the flipped classroom sessions [13].

In the traditional teaching method, the focus is on content delivery and not on individual needs. This restricts the clarification of concepts and doubts for each student. However, in the flipped classroom method individualized differentiated learning was facilitated by the personalized assignments. So, there is need of more facilitators during the flipped session. The time and effort required from the faculties as facilitators will be a challenge in the initial period. However, this exercise once done will be very effective and time saving in the long run [4].

If the entire course material can be appropriately converted into flipped sessions, it will serve as time saving for the students who do not need in class help. Students can focus their efforts on their individual learning need. Thus, they are not left behind by class discussions that go too fast or become bored by class time that is spent covering content they already know. Students need attend class only if they need help beyond what is provided by other learning resources. This can be helpful for the efficient students to cover the course material in a shorter duration and use the additional time for gaining psychomotor skills pertaining to patient care [4].

Flipped classroom can also be converted into e-flipped classroom sessions. We suggest development of app for all subjects which will have pre-recorded videos, ppts, notes etc. so the students will not waste time in surfing on internet to understand concepts and clearing doubts as is done for traditional classroom teaching. This authenticated study material can be customized based on local need of the institution. This will aid learning as well as assessing the medical graduates, especially during unprecedented times like COVID-19, when along with patient care there is an urgent need to focus on education of future physicians as well. This global emergency requires intense and prompt attention from medical educators.

## 7. Conclusion

Active participation of students, sharing of their fundamental knowledge and style of learning during flipped classroom sessions add to learning skills of students. Flipped classroom sessions can be incorporated frequently for different topics and disciplines. It can be modified to be used for online teaching sessions and assessments. Thus, the study concludes that flipped classroom models serves as an advantageous tool and motivating factor for effective learning, understanding and retention of conceptual and factual content.

## Data Availability

All data referred to in the manuscript is available

## Acknowledgements

The authors wish to convey their sincere thanks to the Director, AIIMS, Nagpur and all staff members from the Department of Anatomy, AIIMS, Nagpur. Authors also acknowledge the immense help received from the scholars whose articles are cited and included in references of this manuscript.

